# Saliva as an alternative sample type for detection of pneumococcal carriage in young children

**DOI:** 10.1101/2023.06.26.23288970

**Authors:** Anne L. Wyllie, Nynke Y. Rots, Alienke J. Wijmenga-Monsuur, Marlies A. van Houten, Elisabeth A.M. Sanders, Krzysztof Trzciński

## Abstract

In children, the gold standard for the detection of pneumococcal carriage is conventional culture of a nasopharyngeal swab. Saliva, however, has a history as one of the most sensitive methods for surveillances on pneumococcal colonisation and has recently been shown to improve carriage detection in older age groups. Here, we compared the sensitivity of nasopharyngeal and saliva samples from PCV7-vaccinated 24-month-old children for pneumococcal carriage detection using conventional and molecular diagnostic methods.

Nasopharyngeal and saliva samples were collected from 288 24-month-old children during the autumn/winter, 2012/2013. All samples were first processed by conventional diagnostic techniques. Next, DNA extracted from all plate growth was tested by qPCR for the presence of pneumococcal genes *piaB* and *lytA* and a subset of serotypes.

By culture, 164/288 (57%) nasopharyngeal swabs tested positive for pneumococcus, but detection was not possible from saliva due to abundant polymicrobial growth on culture-plates. Molecular methods increased the number of children pneumococci-positive to 172/288 (60%) when testing culture-enriched saliva and to 212/288 (73%) when testing nasopharyngeal samples. Altogether, by molecular methods 239/288 (83%) infants were positive, with qPCR-based carriage detection of culture-enriched nasopharyngeal swabs significantly detecting more carriers compared to either conventional culture (p<0.001) or qPCR-detection of saliva (p<0.001). However, 27/240 (11%) carriers were positive only in saliva, significantly contributing to the overall number of carriers detected (p<0.01).

While testing nasopharyngeal swabs with qPCR proved most sensitive for pneumococcal detection in infants, saliva sampling could be considered as complementary to provide additional information on carriage and serotypes which may not be detected in the nasopharynx and may be particularly useful in longitudinal studies, requiring repeated sampling events.

## INTRODUCTION

The human nasopharynx is considered the primary niche of *Streptococcus pneumoniae*, with colonisation occasionally progressing to pneumococcal disease. Disease manifestations include respiratory infections such as otitis media or pneumonia, or invasive pneumococcal disease (IPD) such as bacteraemia or sepsis with or without meningitis^1^. Current vaccination strategies are targeted towards the polysaccharide capsule, which is considered the primary pneumococcal virulence factor^2^. With over 100 known capsular types (serotypes)^3^, current vaccination coverage is limited to either a maximum of 20 serotypes included in the conjugated polysaccharide vaccines (PCVs) or 23 serotypes for the polysaccharide vaccine (PPSV23). PCV-vaccination of children (the key transmitters of pneumococcus) not only protects against vaccine-serotype (VT) disease, but also against carriage of VT strains. Moreover, PCVs lead to herd protection in other age groups, most importantly, by reducing VT-disease in older adults^1,4–12^. However, the gap left after VT-eradication from carriage in children has been almost completely filled by emerging non-vaccine serotypes (NVTs)^12–14^. This serotype replacement^10^, is eroding the benefits of vaccination to various extents across all age groups, especially in older adults with increasing serotype replacement in diseases like pneumonia and IPD^6,15,16^.

For any strategy aiming to prevent pneumococcal disease, knowledge on pneumococcus reservoirs in the population is essential and surveillance of carriage can provide data on vaccine effects before any impact can be observed in disease^17^. Pneumococcal carriage detection has been instrumental in monitoring the impact of vaccination against pneumococcal disease. The gold standard method for carriage detection is conventional culture of a nasopharyngeal swab with an additional oropharyngeal swab recommended to improve the sensitivity of detection in adults^18^. Historical records from the pre- antibiotic era however, reported high carriage rates ranging between 39% and 54% across all ages, when oral (saliva) samples were tested with sensitive animal inoculation method^19–21^. This suggested to us that sampling of the oral niche might increase carriage detection^22^. In line with this, we and others have demonstrated the potential of molecular methods for increased pneumococcal detection in older age groups when oropharyngeal swabs^23–26^ or saliva samples were tested alongside nasopharyngeal swabs^22,23^.

Since young children are typically the focus of surveillance on pneumococcal carriage prior to or following updated vaccination strategies, we explored the sensitivity of culture and molecular methods for pneumococcal carriage detection in nasopharyngeal swabs and saliva samples collected from 24-month-olds to investigate whether nasopharyngeal sampling was also under detecting carriage prevalence in this population.

## METHODS AND MATERIALS

### Study design

Nasopharyngeal swabs were collected in a prospective cross-sectional study conducted in the Netherlands during the autumn/winter season of 2012/2013 from 330 PCV7-vaccinated 24-month-old children^14,27^. Detailed descriptions of the study population and primary results for pneumococcal carriage detection in these nasopharyngeal samples were reported on previously^14,27^ and in the current study, were analysed together with saliva samples which were also collected, as described below.

The study was approved by METC Noord-Holland (NL40288.094.12)^14^ and conducted in accordance with the European Statements for Good Clinical Practice and the declaration of Helsinki of the World Health Medical Association. Written informed consent was obtained from all parents.

### Collection of saliva samples

At time of nasopharyngeal sample collection^14^, a saliva sample was also collected from each PCV7-vaccinated 24-month-old child. Prior to sample collection, informed consent was obtained from both parents/caregivers, with saliva collection also approved by METC Noord-Holland (NL40288.094.12)^14^. Briefly, a saliva collection sponge (Oracol®, Malvern Medical Developments, Worcester, United Kingdom) was placed in the front part of the child’s mouth for approximately one minute until saturated with saliva^28^. The wet sponge was then placed into a 5 ml syringe and the stick was withdrawn through the narrow opening, leaving only the sponge inside the syringe. Using the plunger, the sponge was compressed, and the saliva was transferred to 2 ml cryovials prefilled with 0.1 ml of 50% glycerol water solution. Samples were transported to the diagnostic lab on dry ice, stored at -80°C, thawed in batches and 100 µl of saliva cultured on Columbia agar with 7% defibrinated sheep blood and gentamicin 5 mg/l, the medium selective for pneumococcus, as previously described^14,23^. All bacterial growth was harvested from all culture-plates into Brain Heart Infusion Broth (Oxoid, Badhoevedorp, the Netherlands) supplemented with 10% glycerol and stored frozen at -80°C^22^. These samples were considered culture-enriched for pneumococcus. Later, DNA was extracted from 200 _μ_l of culture-enriched saliva samples as previously described^22^.

### Pneumococcal carriage and serotype detection

DNA templates from culture-enriched saliva were first tested for the presence of two pneumococcal-specific genes *piaB*^24,27^ and *lytA*^29^. Samples were classified as positive for pneumococcus when C_*T*_ values for both targeted genes were <40^22,23,25,26^.

Regardless of outcome of *piaB* and *lytA* qPCR-testing, all saliva samples from children were tested in qPCR for the presence of sequences specific for pneumococcal serotypes/serogroups 1, 3, 6A/B/C/D, 7A/F, 8, 9A/N/V, 10A/B, 12A/B/F, 14, 15A/B/C, 19A, 20, 23F, 33A/F/37^30^, 11A/D, 16F, 18B/C and 19F^31^. Samples were classified as positive for pneumococcal serotype/serogroup when C_*T*_ values for targeted genes were <40.

### Statistics

Statistical analyses were conducted using GraphPad Prism v5.0 (GraphPad Software, San Diego, CA, USA). Differences in pneumococcal serotype carriage and serotype detection were tested for using two-way Fisher’s exact tests for 2×2, or Chi-square for 3×2 contingency tables. An estimate was considered statistically significant at *p*<0.05.

## RESULTS

From the 293 24-month-old children previously reported on^27^ for pneumococcal carriage detection in their nasopharyngeal samples, 288 (98%) matching saliva samples were available for inclusion in the current study. Results summarised in Table 1 depict differences in pneumococcal carriage detection between the two sample types and diagnostic methods (culture vs. molecular). Here, we report on the sensitivity of each method, defined as the number of carriage events detected by each method, over the total number of carriage events detected by all methods combined.

**Table 1.** Sensitivity of pneumococcal carriage detection in nasopharyngeal and saliva samples collected from PCV7-vaccinated 24-month-old children using conventional and molecular diagnostic methods.

| **24-month-olds (n=288)<sup>27</sup>** |  |  |
| --- | --- | --- |
| **Detection method** | **Carriage** | **Sensitivity** |
| **Culture** |  |  |
| Nasopharyngeal | 161 (60%) | 0.74 |
| Saliva | ND<sup>a</sup> | 0 |
| **qPCR** |  |  |
| Nasopharyngeal | 187 (65%)<sup>#</sup> | 0.85* |
| Saliva | 155 (54%) | 0.71 |
| **Overall** | **219 (76%)<sup>##</sup>** | **1** |

<sup>a</sup>ND, not detectable by culture due to abundant polymicrobial growth on culture-plate
<sup>#</sup>p<0.05, <sup>##</sup>p<0.001 (Fisher's exact test), significantly more carriers detected by this approach as compared to the gold standard method of culture of nasopharyngeal swab (top row)
\*method significantly more sensitive in carriage detection (p<0.05) than any other tested in the particular study group

### Pneumococcal carriage detected by culture

Overall, culture-detected carriage rates were in line with contemporary rates reported by others^14,32,33^, with a relatively high sensitivity of nasopharyngeal carriage detection (0.74). Isolation of live pneumococci from saliva at the initial culture step was virtually impossible due to abundant polymicrobial growth on plates selective for pneumococcus^22,23^, thus the sensitivity of pneumococcal carriage detection by culturing saliva at the primary diagnostic step was null.

### Pneumococcal carriage detected in culture-enriched samples by qPCR

Testing culture-enriched samples with qPCR significantly increased pneumococcal detection as compared with culture detection. Testing culture-enriched nasopharyngeal samples by the molecular method was the most sensitive method of carriage detection (0.85) and identified significantly more carriers than testing culture-enriched saliva samples with qPCR (187/288, 65% versus 155/288, 54%; *p*<0.001). Nonetheless, saliva significantly contributed to the overall carriage rate detected compared to testing nasopharyngeal swabs alone (219 versus 187, *p*<0.005). Importantly, there was no difference between the prevalence of carriage detected by testing saliva by qPCR compared with the gold standard culture of nasopharyngeal swabs (155 versus 161, *p*=0.68).

Overall, 219/288 (76%) children were positive for pneumococcus when results of all methods applied were combined.

### Effect of sample type and testing method on pneumococcal serotype detection

Since we report here for the first time on pneumococcal carriage detection using saliva of children, we compared this data to the previously described nasopharyngeal serotype carriage data^27^, detected by either the recommended culture-based approach or by testing culture-enriched nasopharyngeal samples using molecular methods (Table S1).

As compared to culture-based pneumococcal detection in nasopharyngeal samples, application of molecular methods to saliva detected significantly more carriers of serotypes 11A/D (19/161, 12% versus 33/155, 21%; *p*=0.033), 19A (25/161, 16% versus 39/155, 25%; *p*=0.036) and PCV13-VTs overall (35/161, 22% versus 50/155, 32%; *p*=0.042) (Figure 1A). However, when both sample types were tested by qPCR, there were no differences in the carriage frequencies of serotypes between nasopharyngeal or saliva samples (Figure 1B). Moreover, there was a strong correlation between the frequency of a serotype detected in nasopharyngeal samples by culture and its frequency of detection in saliva by qPCR (rho=0.845; *p*<0.001), which was even stronger when results of carriage detection with molecular methods for nasopharyngeal and saliva samples were compared (rho=0.849, *p*<0.0001).

**Figure 1.**
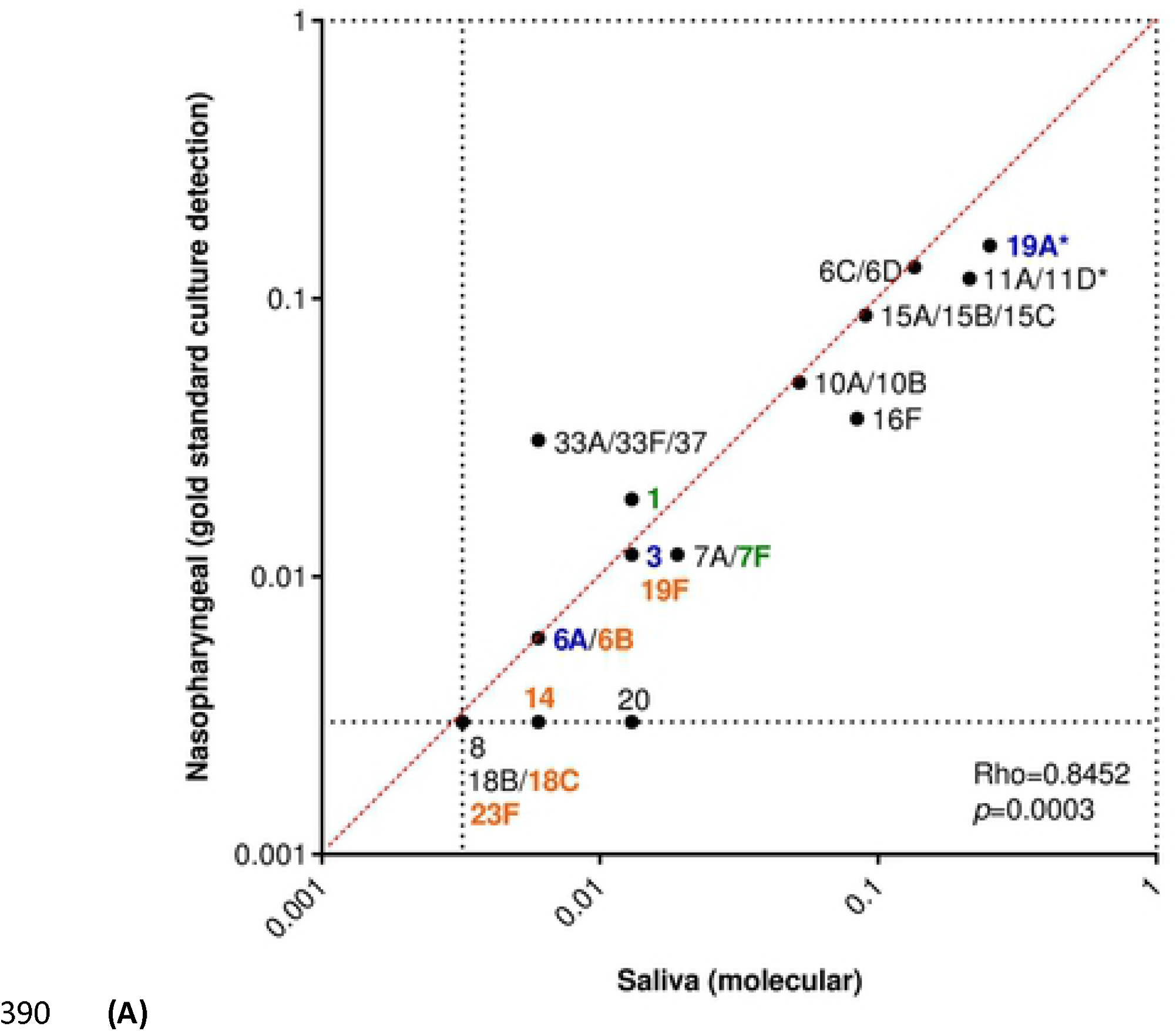
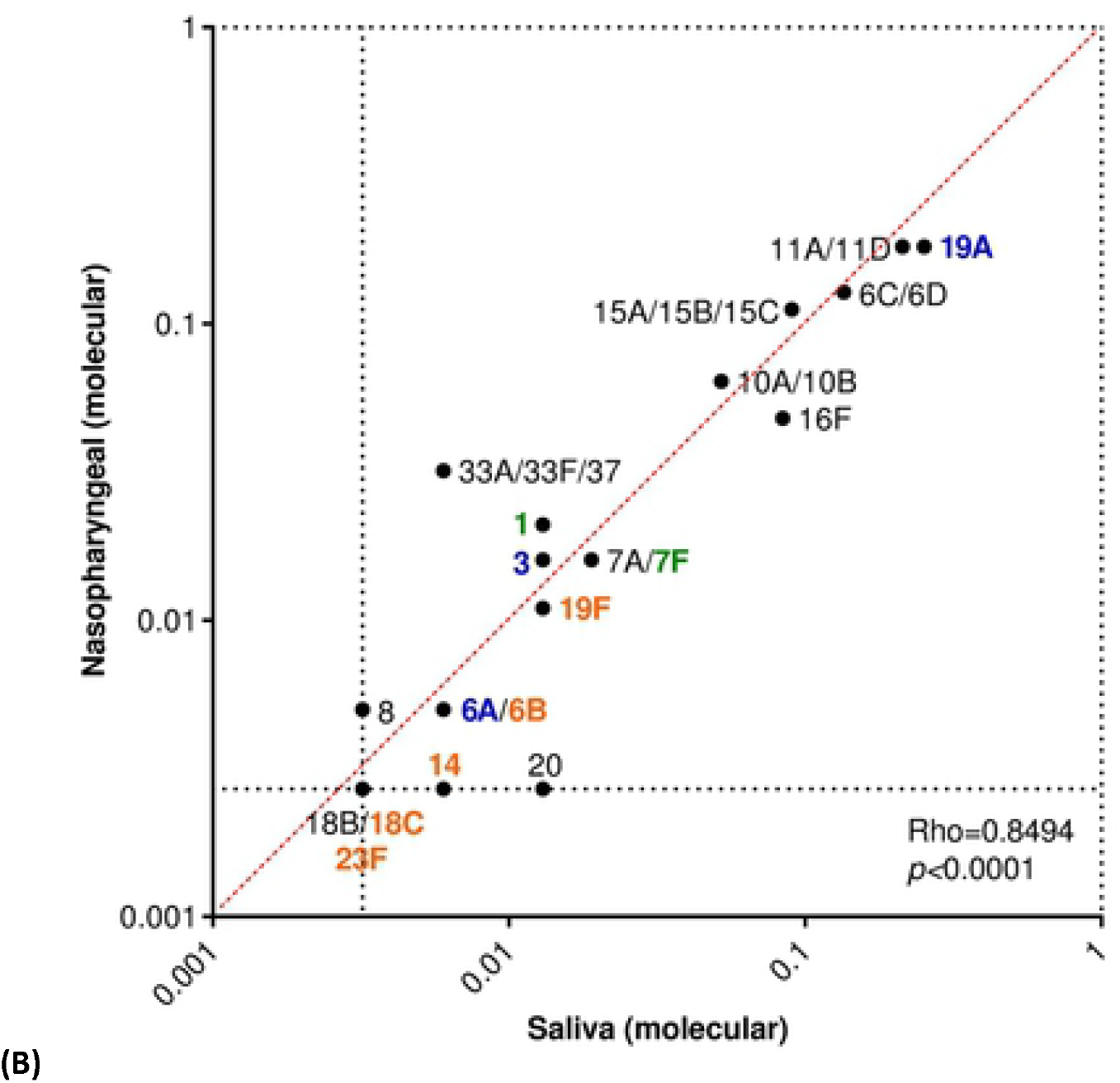
Carriage frequency of pneumococcal serotypes detected in nasopharyngeal and saliva samples from 24-month-old children, when tested by conventional culture or molecular methods. Graphs depict the correlation between the overall carriage frequency of serotypes (from the subset targeted by qPCR), as detected by (A) culture or (B) molecular methods applied to nasopharyngeal swabs compared to the overall carriage frequency of the corresponding serotype when saliva samples were tested by qPCR. The frequency of carriage was calculated for each serotype by the total number of samples testing positive for particular serotype by either the Quellung or molecular methods, over the total number of pneumococcal carriers detected for each study group. Serotypes not detected by only one of the methods were assigned a value of 0.5 x the fraction representing a single carrier. Serotypes not detected by both methods were excluded from correlation calculations. Font colour indicates serotypes targeted by PCV7 (orange), PCV10 (green), PCV13 (blue) or NVTs (black). Asterisks depict serotypes which differed significantly (p<0.05) in frequency of carriage between sample types.

With similar rates of overall pneumococcal carriage detection in both nasopharyngeal and saliva samples collected from children, we were interested in the benefit of each sample type for its contribution to serotype detection. Therefore, to investigate the additive effect of samples obtained from different niches on serotype detection in children, we re-analysed the serotyping data, excluding children who were positive for the same serotype in both niches. As compared to culture detection in nasopharyngeal samples, testing saliva with qPCR detected significantly higher carriage of serotypes 16F (0/161, 0% versus 5/155, 3%; *p*=0.027) and 19A (5/161, 3% versus 16/155, 29%; *p*=0.012; respectively) (Figure 2A). When both sample types were tested by qPCR, there were no significant differences in additional serotypes detected by either sample type (Figure 2B).

**Figure 2.**
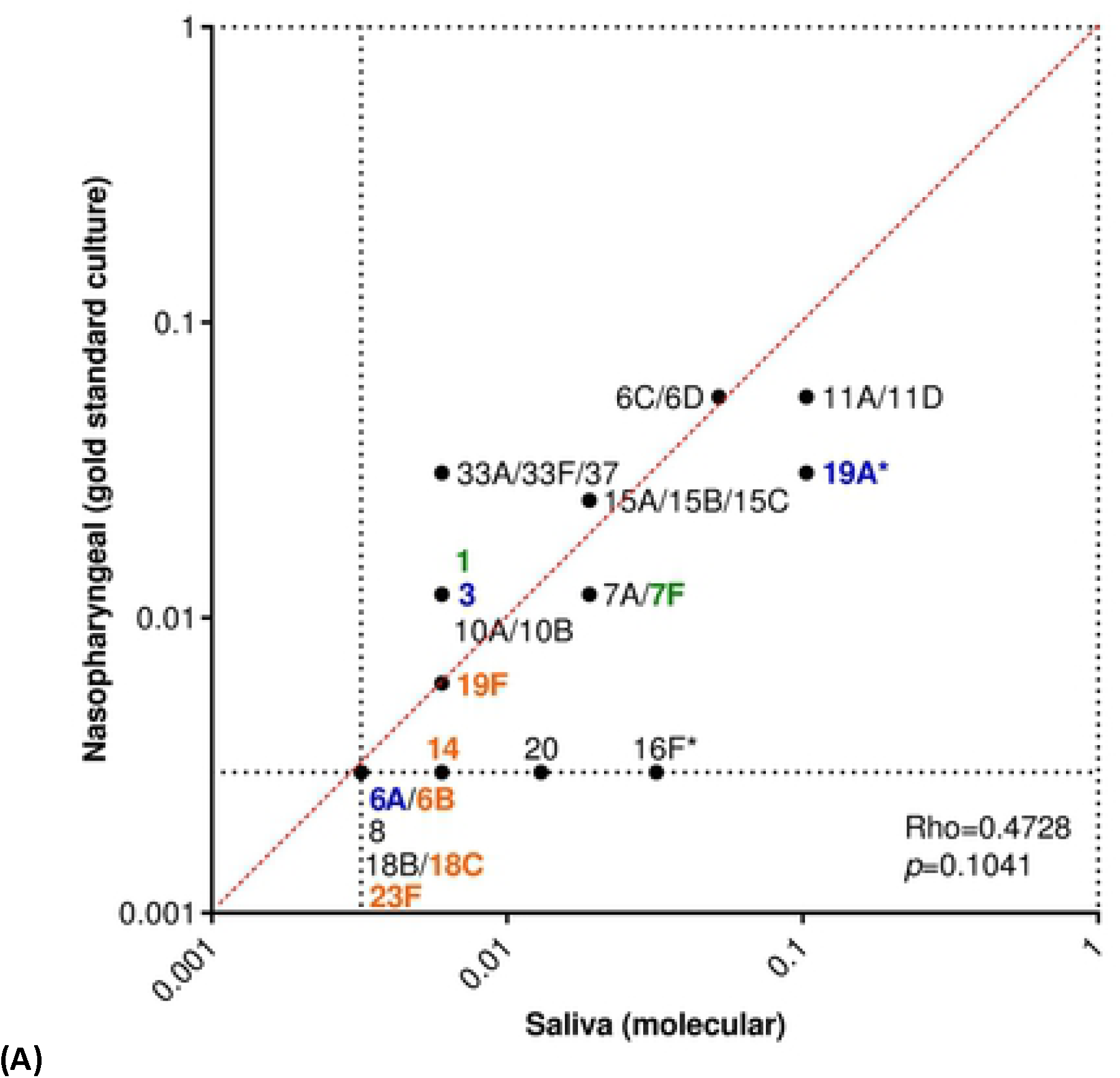
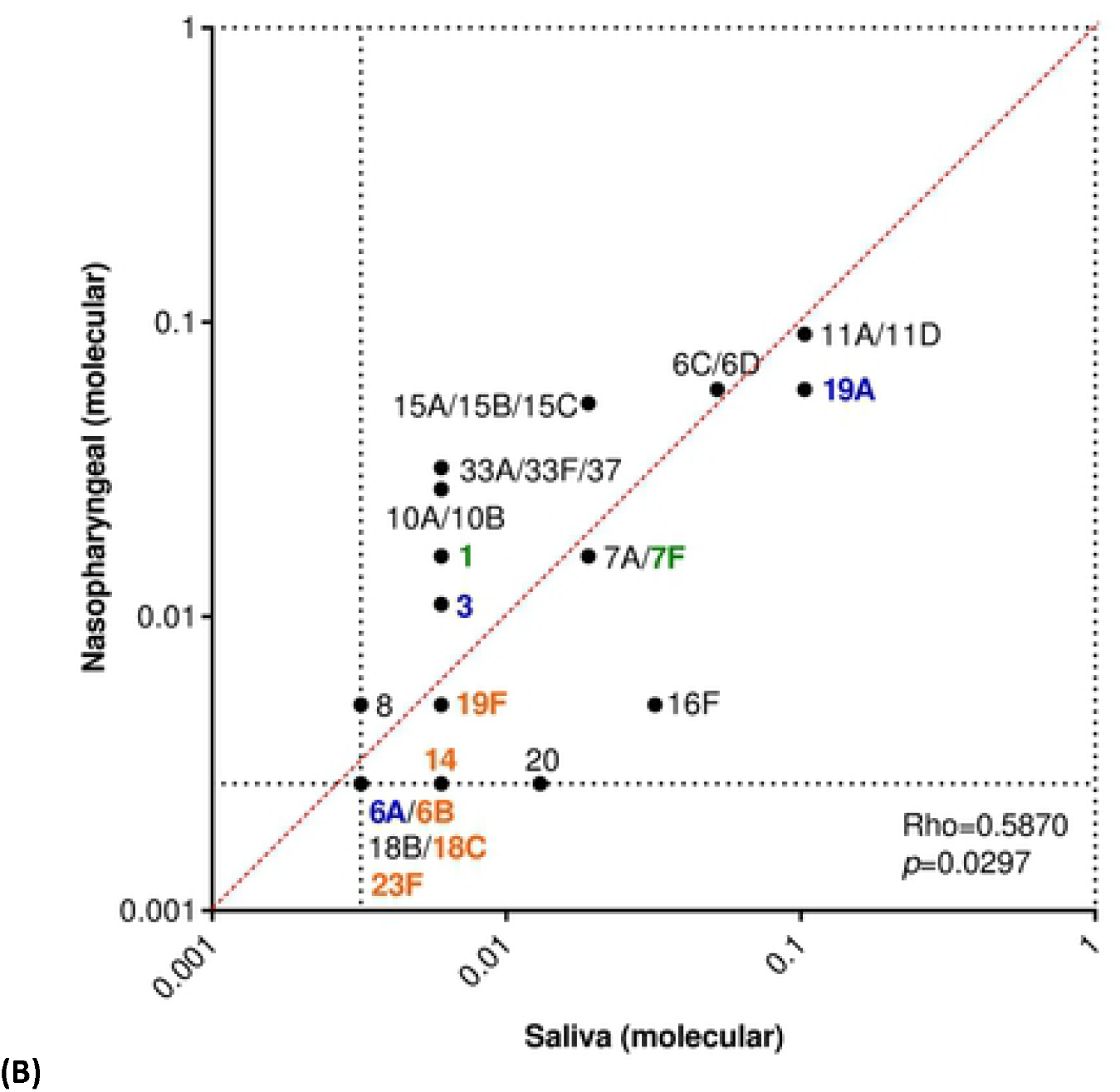
Investigating the additive effect of testing samples obtained from different niches on the frequency of pneumococcal serotypes detected in 24-month-old children. Graphs depict serotype carriage frequency in pneumococcal carriers who tested positive for a specific serotype in only one sample type; individuals who were positive for the same serotype in both sample types were excluded from analysis. The frequency of carriage was calculated for each serotype by the total number of samples testing positive for that particular serotype by either the Quellung or molecular methods, over the total number of pneumococcal carriers detected by that method. When a serotype was detected by only one of the methods, for the method it was not detected by, it was assigned a value of 0.5 x the fraction representing a single carrier. Serotypes not detected by both methods were excluded from correlation calculations. While (A) significantly higher carriage frequencies of serotypes 16F and 19A were detected when saliva was tested by qPCR as compared to the gold standard culture-based method (*p*<0.05), (B) no serotype was detected more frequently by either sample type when samples were tested by qPCR. Font colour indicates serotypes targeted by PCV7 (orange), PCV10 (green), PCV13 (blue) or NVTs (black). Asterisks depict serotypes which differed significantly (*p*<0.05) in frequency of carriage between sample types.

## DISCUSSION

The current culture-based recommendation for detecting pneumococcal carriage is becoming more frequently challenged^18,23–25,34–36^. While culture-independent methods improve the sensitivity of carriage detection in nasopharyngeal samples from both children^30,37,38^ and adults^23–26,39^, we have also demonstrated improved sensitivity when these methods are applied to alternative respiratory samples from adults^23–25^. Therefore, in the current study, we conducted a direct comparison of nasopharyngeal swab and saliva samples collected from 24-month-old children, for their sensitivity for pneumococcal detection when processed by culture and molecular methods; nasopharyngeal sampling proved superior in children.

While nasopharyngeal swabs were optimal for overall detection of pneumococcal carriage in children, saliva significantly contributed to the overall carriage rate (219 total carriers detected versus 187 carriers detected by nasopharyngeal swabs alone, *p*<0.005, and 155 by saliva alone). For serotype carriage detection however, neither nasopharyngeal nor saliva samples were superior when all samples were processed by the molecular method. This suggests to us that results for serotype distribution generated using saliva are equally representative to results generated with the current gold standard method of conventional culture of nasopharyngeal swabs in children. It should be noted that culture-enrichment can enhance carriage detection in low density carriers or of secondary (or lesser) serotypes co-carried in a sample when serotype surveillance is being considered. The differences between serotypes detected in saliva compared to serotypes cultured from nasopharyngeal swabs were likely due to failure of colony picking during culture to detect co-carriage of less abundant strain(s)^40^. Importantly, overall serotype detection in saliva samples correlated well with qPCR-detection in nasopharyngeal swabs. Hence, molecular testing of saliva samples holds potential for improving surveillance of pneumococcal carriage, providing additional insight into pneumococcal carriage compared to sampling the nasopharynx alone or providing a means to reduce the burden of study protocols through simplified sample collection. Additionally, with little-to-no discomfort from non-intrusive collection, saliva sampling is generally better tolerated compared to nasopharyngeal swabbing encouraging greater adherence to sampling routines^23^, while reducing the number of participants or samples lost due to testing aversion and refusing of sample collection^23^. This makes saliva particularly suitable for longitudinal studies.

The vast majority of epidemiological surveillance studies of pneumococcal carriage are descriptive, based on qualitative results of carriage detection and focus on serotype distribution. They are usually solely aimed at monitoring the disappearance of VTs and the emergence of NVTs. Future vaccines may transcend pneumococcal serotypes, with vaccines protecting against all pneumococci, independent of capsular polysaccharide expressed. In this instance, while determining serotype-specific carriage may be only a secondary interest, accurate measures of overall pneumococcus in its ecological niche (the presence and density of all serotypes combined) will remain essential for monitoring vaccine effects and establishing study endpoints. More complex studies, with repeated sampling events, are required for better understanding of carriage dynamics (rates of acquisition and clearance, episode length) in both carriage and disease and in children versus adult populations.

With the increasing availability of more sensitive detection methods, it is again being reported that as in the early studies, pneumococcal carriage can be long-lasting, and that co-carriage of multiple serotypes is common. Longitudinal carriage of serotypes increases the risk of transmission, but also provides opportunities for intra- and inter-species genetic recombination. While findings from this and previous studies suggest to us that no single sample type should be considered as universally superior for pneumococcal carriage detection across all age groups, collecting saliva from children – whether alone or in addition to nasopharyngeal swabs - could be considered as a quick, non-invasive option for enhanced detection with greater serotype carriage. If strain isolation is not of critical importance, the two sample types could be merged and tested as one. Moreover, sampling saliva may have an even greater benefit in broader studies for its potential to also be tested for other upper respiratory tract commensals or pathogens, such as meningococci^41,42^, as well as for antibody responses.

## Data Availability

All data produced in the present study are available upon reasonable request to the authors

## DECLARATIONS

### Conflict of Interest

ALW has received consulting and/or advisory board fees from Pfizer, RADx, Diasorin, PPS Health, Co-Diagnostics, Filtration Group, and Global Diagnostic Systems for work unrelated to this project, and is Principal Investigator on research grants with Pfizer, Merck, Flambeau Diagnostics, Tempus Labs, and The Rockefeller Foundation to Yale University. MAvH declares to have received research grants from Pfizer. EAMS declares to have received research grants from Pfizer and GSK and fees paid to the institution for advisory boards and participation in independent data monitoring committees for Pfizer and GSK. KT declares to have received research grants from Pfizer and GSK and fees for advisory boards for Pfizer, all paid to the home institution. All other authors report no potential conflicts.

### Funding

The work was funded by the Dutch Ministry of Health. Unrestricted grant support for molecular microbiology was provided by Pfizer (the Netherlands), through investigator-initiated research grants (WS2312079 and WS2312119 to EAMS and KT).

## Acknowledgements

We gratefully acknowledge the participating families for their time and commitment to the study. We thank all members of the research team of the Spaarne Gasthuis, the laboratory staff of Regional Laboratory of Public Health, Haarlem, the Netherlands and the cooperating institutes for their dedication and work which made this project possible. In particular, we would like to thank Astrid Bosch for assistance with sample collection and Lidewij Rümke and Jody van Engelsdorp Gastelaars for laboratory assistance.

## Authors’ contributions

ALW, NR, EAMS and KT had the idea and initiated the study. AJWM, NR, EAMS and KT wrote the study protocols. ALW, AJWM and MAvH managed the study and collected the data. ALW was responsible for and performed the assays. ALW and KT analysed and interpreted the data. ALW, EAMS and KT drafted the manuscript. All authors amended and commented on the final manuscript.

## Notes

### Author Declarations

The study was approved by METC Noord-Holland (NL40288.094.12) and conducted in accordance with the European Statements for Good Clinical Practice and the declaration of Helsinki of the World Health Medical Association. Written informed consent was obtained from all parents.

